# Burden and Seasonal Variations of Cutaneous Leishmaniasis in Afghanistan During 2024

**DOI:** 10.64898/2026.08.10.26360070

**Authors:** Bilal Ahmad Rahimi, Wais Mohammad Lali, Ahmad Jawad Sherzad, Walter R. Taylor

**Author notes:** **Corresponding Author,** Bilal Ahmad Rahimi, MD, DTM&H, MCTM(TP), PhD, E-mail address. Designations: Department of Public Health, Faculty of Medicine, Afghan International Islamic University, Kabul, Afghanistan. Department of Pediatrics, Faculty of Medicine, Kandahar University, Kandahar, Afghanistan. Postal address: Department of Public Health, Faculty of Medicine, Afghan International Islamic University, Dar- ul-Aman, Kabul, Afghanistan.

## Abstract

**Background:** Cutaneous leishmaniasis (CL) remains a major neglected tropical disease in Afghanistan; however, recent nationwide evidence on its geographical and temporal distribution is limited. This study assessed the reported burden, spatial distribution and seasonal variation of cutaneous leishmaniasis across Afghanistan during 2024.

**Methods:** A nationwide retrospective ecological study was conducted using aggregated routine surveillance data obtained from the National Malaria and Other Vector-Borne Diseases Program. All cases reported from government health facilities across Afghanistan’s 34 provinces between 1 January and 31 December 2024 were included. Reported incidence rates were calculated per 10,000 population using national and subnational population estimates. Cases were analyzed by surveillance classification, province, city, rural district, month, and season.

**Results:** A total of 87,245 cases were reported during 2024, corresponding to an overall incidence of 25.1/10,000 population. Anthroponotic and zoonotic CL accounted for 80,202 (91.9%) and 7,043 (8.1%) cases, respectively. Jowzjan recorded the highest provincial incidence (105.0/10,000), while Kabul and Herat contributed the largest absolute numbers of cases.

Shiberghan had the highest urban incidence, whereas Panjwai, Ghoryan and Hazrat-e-Sultan recorded exceptionally high rural rates. May had the highest monthly burden, while August had the lowest. Autumn accounted for the largest seasonal proportion (27.92%), whereas summer had the lowest burden (16.98%). Considerable incompleteness in district-level reporting was observed.

**Conclusion:** CL constituted a substantial but highly concentrated public health burden in Afghanistan during 2024. Strengthened surveillance, improved diagnostic confirmation and geographically targeted case-management and vector-control interventions are particularly needed in identified high-burden provinces, cities and rural districts.

## Introduction

Cutaneous leishmaniasis (CL) is a neglected tropical disease caused by protozoan parasites of the genus Leishmania and transmitted through the bite of infected female phlebotomine sandflies [1,2]. The disease is endemic in more than 90 countries across tropical and subtropical regions, with an estimated 0.7–1.2 million new cases occurring globally each year [3,4]. CL presents with a range of cutaneous manifestations, including papules, nodules, plaques and ulcers, which may heal with permanent scarring and disfigurement. Although the disease is rarely life-threatening, its visible lesions can cause considerable psychological distress, social stigma, discrimination and economic hardship, particularly among women and children [5–8].

CL remains an important public health problem across the Eastern Mediterranean Region and neighbouring areas, including Morocco, Algeria, Tunisia, Libya, Iran, Pakistan and Saudi Arabia [1,5]. Epidemiological studies from these countries demonstrate substantial variation in the geographical distribution, seasonality and population groups affected by the disease. In Libya, CL has been associated with agricultural and outdoor occupations, while studies from several provinces of Iran have reported considerable case burdens and variation according to age, sex,

lesion location and season [9–13]. Molecular evidence from north-eastern Iran has also demonstrated the coexistence of *Leishmania tropica* and *Leishmania major* in areas previously considered to support a single transmission pattern, illustrating the dynamic nature of CL epidemiology [14].

A similarly heterogeneous pattern has been reported in Pakistan, particularly in Khyber Pakhtunkhwa Province, which shares a border with Afghanistan. Recent investigations in Bajaur, Malakand and Dir Lower documented a substantial burden among children and young people, with lesions frequently occurring on the face and other exposed parts of the body [1]. Cases reported in Karachi have also been linked to travel from endemic areas, suggesting that population movement may contribute to the introduction of CL into urban settings [15]. In Saudi Arabia, surveillance from Asir Province likewise showed geographical clustering and a high burden among children [16]. Collectively, these findings indicate that CL transmission is highly focal and shaped by interactions among ecological conditions, vector activity, human mobility, living environments and access to healthcare.

Afghanistan has historically been recognised as one of the countries with the highest burden of CL worldwide [3]. Both principal epidemiological forms have been reported: anthroponotic cutaneous leishmaniasis (ACL), commonly associated with *L. tropica* and predominantly reported in urban settings, and zoonotic cutaneous leishmaniasis (ZCL), commonly associated with *L. major* and more frequently reported in rural and semi-arid areas [2]. Historical studies demonstrated particularly intense transmission in Kabul. A survey conducted in 2002 found evidence of previous infection, based on characteristic scars, in more than two-thirds of the population in some districts, while approximately 67,500 active cases were estimated in Kabul during 2002–2003 [17].

More recent evidence indicates that CL remains widely distributed in Afghanistan, but contemporary national-level analyses are limited. Continuing conflict, large-scale population displacement, fragile health infrastructure and climatic variability may influence both disease transmission and the ability of the health system to detect and report cases [18]. A cross-sectional study conducted in Kabul during 2020–2021 reported CL among 11.4% of suspected patients, with children and young adults commonly affected and lesions occurring predominantly on the face and hands [19]. An analysis of cases reported from eastern Afghanistan between 2017 and 2022 identified Nangarhar as the most affected province within that region and found higher case detection during autumn and winter [20]. Nevertheless, recent evidence describing the reported burden of CL across all 34 provinces, together with its urban, rural and temporal distribution, remains scarce.

Temporal variation is an important feature of CL epidemiology. The activity and abundance of sandfly vectors are influenced by temperature, humidity and rainfall, while the timing of clinical presentation may also reflect the incubation period, delays in care-seeking and surveillance practices [5,11]. Studies from Iran have frequently reported higher case numbers during autumn, potentially reflecting infections acquired during periods of greater sandfly activity in late spring and summer and becoming clinically apparent after an incubation period of several weeks or months [13,21]. In contrast, studies from Pakistan have reported higher numbers during spring and summer, indicating that temporal patterns may differ between epidemiological settings [1]. Understanding the monthly distribution of reported cases in Afghanistan is therefore important for planning surveillance, preparing diagnostic and treatment services, timing vector-control activities and allocating limited public health resources.

Despite the long-recognized burden of cutaneous leishmaniasis in Afghanistan, recent nationwide evidence remains limited. Existing studies have largely focused on individual cities, provinces or selected regions and have not simultaneously examined provincial, urban, rural- district and temporal patterns across all 34 provinces. Consequently, the geographical concentration of reported disease, the relative contribution of high-burden areas to the national caseload and the monthly and seasonal distribution of reported cases remain insufficiently characterized. This evidence gap limits the ability of national programs to identify priority areas, allocate diagnostic and treatment resources efficiently, and design geographically targeted surveillance and control interventions.

The present study addresses this gap by providing a recent nationwide analysis of routinely reported cutaneous leishmaniasis across all 34 provinces of Afghanistan, integrating absolute case burden with population-based incidence estimates and examining variation at provincial, urban, rural-district, monthly and seasonal levels. By distinguishing areas with large service burdens from those with exceptionally high incidence, the study provides a more detailed epidemiological baseline for prioritizing surveillance, case management and locally adapted control measures. Accordingly, this study aimed to describe the reported burden and geographical distribution of cutaneous leishmaniasis across Afghanistan during 2024.

Specifically, it estimated national and subnational reported incidence rates and examined the distribution of reported cases across provinces, urban centers, rural districts, months and seasons.

## Materials and Methods

### Study design and setting

A nationwide retrospective ecological study was conducted using routinely collected surveillance data on cutaneous leishmaniasis reported in Afghanistan between 1 January and 31 December 2024. The study covered all 34 provinces and examined the reported distribution of cases at national, provincial, urban and rural-district levels. Monthly case counts were also analyzed to describe temporal variation during the study period.

Afghanistan has substantial geographical, climatic and demographic diversity, encompassing densely populated urban centers and extensive rural, mountainous and semi-arid areas. These settings may provide different ecological conditions for anthroponotic and zoonotic transmission. Because the source data were aggregated by geographical reporting unit provinces, cities and districts, rather than individual patients, these constituted the units of analysis.

### Data source and surveillance system

Aggregated surveillance data were obtained from the National Malaria and Other Vector-Borne Diseases Program of the Afghanistan Ministry of Public Health. The source dataset contained monthly numbers of reported cutaneous leishmaniasis cases for 2024, organized by province, city or urban reporting area, and district.

The surveillance data originated from routine reporting by government health facilities and were subsequently compiled by the national program. The dataset provided to the investigators contained no names, identification numbers, residential addresses or other personal identifiers. Individual-level information on demographic characteristics, clinical presentation, diagnostic procedures, treatment and outcomes was not available.

### Study population and eligibility

The study population comprised all cutaneous leishmaniasis cases recorded in the national surveillance dataset during 2024. Records from government health facilities in all 34 provinces were eligible for inclusion.

A record was included when it contained a recognized geographical reporting unit and a documented number of cutaneous leishmaniasis cases during the study period. Records outside the 2024 reporting period and entries that could not be assigned to a recognized province, city or district were excluded from the relevant geographical analysis.

Because the dataset was aggregated, individual patients could not be linked across facilities or reporting periods. The analysis therefore relied on the aggregation and de-duplication procedures applied by the national surveillance program before the dataset was released for analysis.

### Case definition and classification

Cases were identified according to the routine diagnostic and reporting procedures used by the National Malaria and Other Vector-Borne Diseases Program. The source dataset classified reported cases as anthroponotic cutaneous leishmaniasis or zoonotic cutaneous leishmaniasis. The aggregated dataset did not specify the diagnostic method used for each reported case or the proportion confirmed by microscopy, culture, molecular testing or another laboratory procedure. Accordingly, the terms anthroponotic cutaneous leishmaniasis and zoonotic cutaneous leishmaniasis refer to program-defined surveillance classifications and should not be interpreted as species-level confirmation of *Leishmania tropica* or *Leishmania major* in every patient.

### Variables and operational definitions

The primary outcome was the number of cutaneous leishmaniasis cases reported during 2024. The analysis included the following variables:

- Province
- City or urban reporting area
- Rural district
- Month of reporting
- Surveillance classification as anthroponotic or zoonotic cutaneous leishmaniasis
- Estimated population of the relevant geographical area
- Annual reported incidence per 10,000 population
- Monthly and seasonal distribution of reported cases

The national reported burden was defined as the total number of cases recorded in the surveillance dataset during the study year. The contribution of each province to the national burden was calculated as the percentage of all nationally reported cases originating from that province.

Urban analyses were based on cities or urban reporting areas identified in the source dataset and their corresponding population estimates. Rural analyses were conducted at district level. Both absolute case counts and population-based incidence rates were retained because highly populated areas may contribute a large proportion of the national burden despite having lower incidence rates than smaller geographical units.

### Population data and denominators

National and subnational population estimates were obtained from the 2024–2025 population estimates of the Afghanistan National Statistics and Information Authority. The national population denominator used in the analysis was 34,195,527.

Provincial incidence rates were calculated using the estimated population of each province. Urban rates were calculated using the corresponding population estimate for each city or urban reporting area, while district-level rates were calculated using the population estimate of the respective district.

The source workbook provided separate program-defined population denominators of 32,642,026 for anthroponotic cutaneous leishmaniasis and 1,553,501 for zoonotic cutaneous leishmaniasis. These denominators together represented the national population used in the analysis. Because the geographical derivation of the two program-defined denominators was not fully documented in the source workbook, ACL- and ZCL-specific rates were interpreted as surveillance-based estimates rather than definitive species-specific incidence measures.

### Data management and quality assessment

The original dataset was organized and reviewed in Microsoft Excel 2021. Geographical names were standardized to reduce inconsistencies in the spelling of provinces, cities and districts.

Province subtotal rows were identified and excluded from district-level calculations to prevent double counting.

Duplicate province–district combinations were examined and consolidated where appropriate. Two additional duplicate records contained no reported monthly cases; their consolidation did not alter the national total. Monthly values were reviewed for negative, non-numeric or implausible entries, and all recorded case values were non-negative integers.

Annual district totals were recalculated by summing monthly case counts. Provincial totals were verified against the sum of the corresponding geographical records. The zoonotic cutaneous leishmaniasis analysis was reconstructed from all 14 Balkh reporting areas, including Mazar-e-Sharif, and the district-level totals were verified against the program-defined total of 7,043 cases and a population denominator of 1,553,501.

Blank monthly cells were retained as indicators of incomplete or unavailable reporting during data-quality assessment. However, where required to reproduce annual totals reported in the source workbook, blank monthly cells were treated computationally as zero contributions to the sum. Because the dataset did not reliably distinguish true zero reports from missing or non-submitted reports, zero and very low values were interpreted cautiously. No statistical imputation was undertaken.

Data completeness was assessed by calculating the number and proportion of blank district– month cells. The cleaned dataset contained 363 district-level records and 4,356 district–month cells.

### Statistical analysis

Descriptive analyses were performed using IBM SPSS Statistics, version 26. Data cleaning, reconstruction of derived tables and independent verification of incidence estimates and confidence intervals were undertaken using Python.

Frequencies and percentages were used to summarize reported cases and the proportional contribution of geographical units to the national burden. Annual reported incidence rates were calculated as:

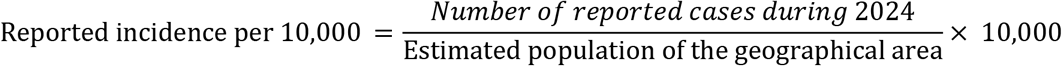

The term reported incidence was used because case numerators originated from routine passive surveillance rather than active population-based case detection.

Exact Poisson 95% confidence intervals were calculated for incidence rates using observed case counts and their corresponding population denominators. Provinces, urban areas and rural districts were ranked according to reported incidence, while absolute case counts were presented separately to distinguish population-based transmission intensity from total service burden.

Monthly counts were arranged chronologically from January to December. For descriptive seasonal aggregation, months were grouped as follows:

- Spring: March–May
- Summer: June–August
- Autumn: September–November
- Winter: December–February

Winter therefore combined January, February, and December 2024 and did not represent one continuous meteorological winter. Seasonal totals were calculated by summing the three months assigned to each season, and the mean monthly count was calculated for each seasonal group.

Because the dataset covered only one calendar year, monthly and seasonal analyses were interpreted as descriptive temporal variation during 2024 rather than as evidence of a stable or recurring seasonal pattern. No inferential tests were conducted to compare geographical areas or seasons, and no causal or statistically significant associations were inferred.

### Geographical presentation

The geographical distribution of reported cutaneous leishmaniasis was examined at provincial, urban and rural-district levels. Provincial results were presented using absolute case counts, reported incidence rates and graphical rankings. For epidemiological mapping, provinces should be grouped according to predefined reported-incidence categories in a choropleth map. The final map should display provincial boundaries, a legend, a north arrow, a scale bar and clear attribution of population and surveillance data sources.

### Ethical considerations

This study involved secondary analysis of aggregated routine public health surveillance data. No patients were contacted, no biological specimens were collected, and the investigators did not have access to personal identifiers. Individual informed consent was therefore not applicable.

Administrative permission to access and analyze the national surveillance data was obtained from the Afghanistan Ministry of Public Health through the National Malaria and Other Vector-Borne Diseases Program. This manuscript states that no primary data or samples were collected and that permission for use of the national data was obtained from the Ministry of Public Health. The data were used exclusively for epidemiological analysis and were handled in aggregated form. No ethics approval or exemption reference number was reported in the source manuscript; therefore, no approval number should be added unless an official letter has been issued by the responsible institutional ethics committee.

## Results

### National burden of cutaneous leishmaniasis

A total of 87,245 cutaneous leishmaniasis (CL) cases were reported across Afghanistan during 2024, corresponding to an overall reported incidence of 25.51 cases per 10,000 population (95% CI 25.34–25.68). Anthroponotic cutaneous leishmaniasis (ACL) accounted for 80,202 cases (91.93%), while 7,043 cases (8.07%) were classified as zoonotic cutaneous leishmaniasis (ZCL). Using the program-defined population denominators provided in the source dataset, the reported incidence was 24.57 per 10,000 population for ACL (95% CI 24.40–24.74) and 45.34 per 10,000 for ZCL (95% CI 44.28–46.41) (Table 1). These classifications represent the categories recorded in the surveillance system and do not necessarily indicate species-level laboratory confirmation for every case.

**Table 1.**
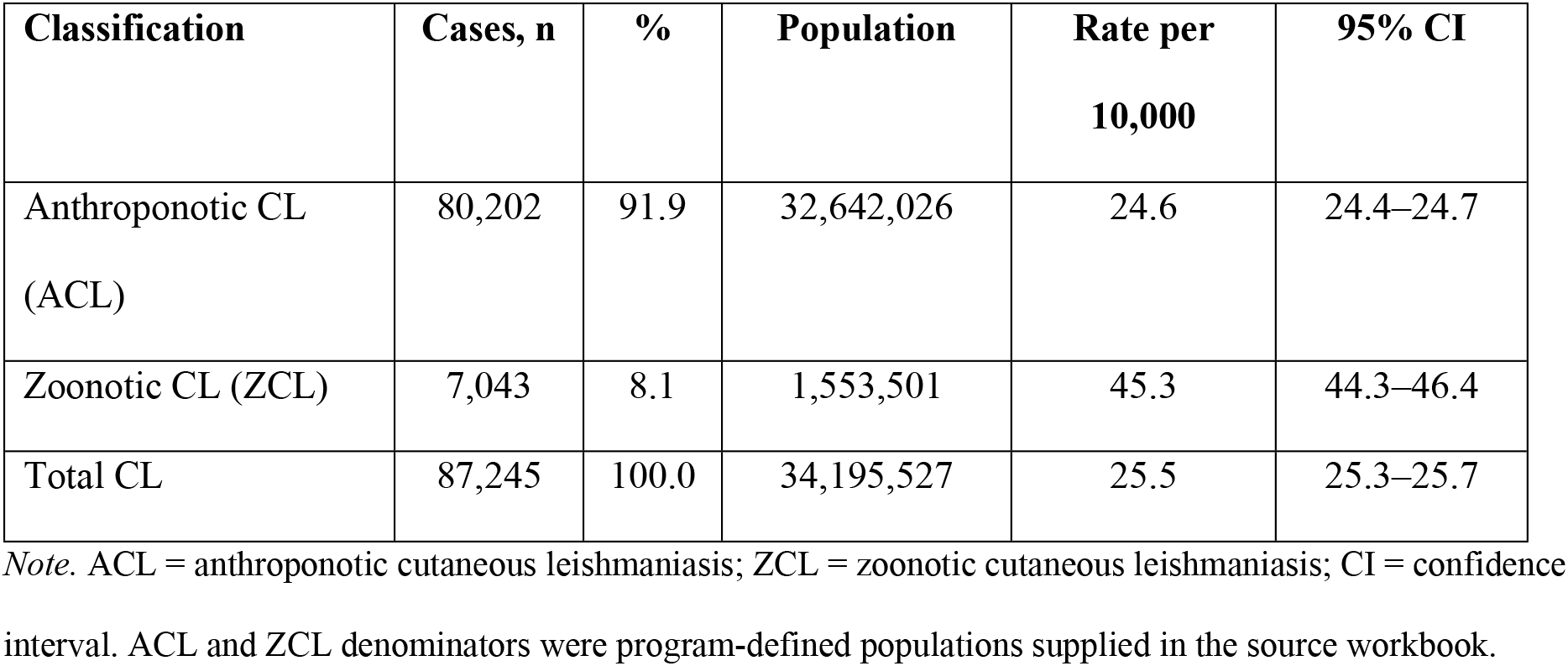
National reported burden and incidence by surveillance classification.

| Classification | Cases, n | % | Population | Rate per<br>10,000 | 95% CI |
| --- | --- | --- | --- | --- | --- |
| Anthroponotic CL<br>(ACL) | 80,202 | 91.9 | 32,642,026 | 24.6 | 24.4–24.7 |
| Zoonotic CL (ZCL) | 7,043 | 8.1 | 1,553,501 | 45.3 | 44.3–46.4 |
| Total CL | 87,245 | 100.0 | 34,195,527 | 25.5 | 25.3–25.7 |

*Note.* ACL = anthroponotic cutaneous leishmaniasis; ZCL = zoonotic cutaneous leishmaniasis; CI = confidence interval. ACL and ZCL denominators were program-defined populations supplied in the source workbook.

### Geographical distribution by province

The reported burden of CL varied markedly across the 34 provinces. Jowzjan had the highest provincial incidence, at 104.99 cases per 10,000 population (95% CI 102.51–107.52), followed by Herat at 62.58 per 10,000 (95% CI 61.57–63.60) and Kandahar at 49.45 per 10,000 (95% CI 48.34–50.57). Other provinces with relatively high reported rates included Laghman, Khost, Kapisa, Logar, Nangarhar and Balkh (Table 2, Figure 1).

**Figure 1.**
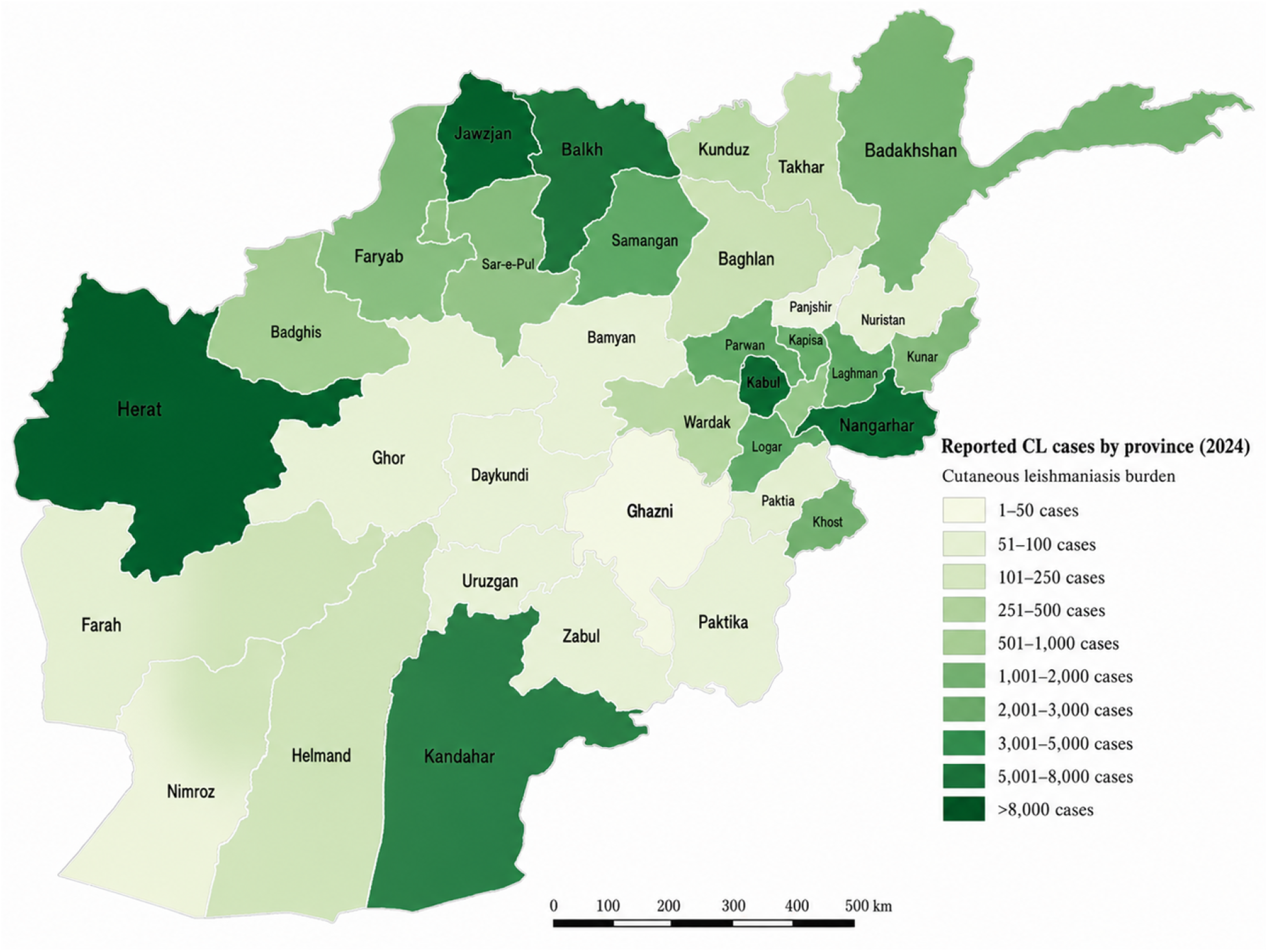
Provincial distribution of reported cutaneous leishmaniasis incidence per 10,000 population in Afghanistan, 2024. The Map Was Produced Using Geographic Information System (GIS) Software.

**Table 2.** Provincial reported cutaneous leishmaniasis burden and incidence, ranked by incidence.

| Rank | Province | Cases, n | % of national<br>cases | Population | Rate per<br>10,000 | 95% CI |
| --- | --- | --- | --- | --- | --- | --- |
| 1 | Jowzjan | 6,812 | 7.81 | 648,804 | 104.99 | 102.51–107.52 |
| 2 | Herat | 14,598 | 16.73 | 2,332,654 | 62.58 | 61.57–63.60 |
| 3 | Kandahar | 7,579 | 8.69 | 1,532,662 | 49.45 | 48.34–50.58 |
| 4 | Laghman | 2,586 | 2.96 | 528,879 | 48.90 | 47.03–50.82 |
| 5 | Khost | 3,211 | 3.68 | 682,333 | 47.06 | 45.45–48.72 |
| 6 | Logar | 2,177 | 2.50 | 465,698 | 46.75 | 44.80–48.75 |
| 7 | Kapisa | 2,442 | 2.80 | 523,201 | 46.67 | 44.84–48.56 |
| 8 | Nangarhar | 8,408 | 9.64 | 1,840,831 | 45.68 | 44.70–46.66 |
| 9 | Balkh | 7,043 | 8.07 | 1,560,365 | 45.14 | 44.09–46.20 |
| 10 | Samangan | 2,152 | 2.47 | 552,763 | 38.93 | 37.30–40.61 |
| 11 | Kunar | 1,895 | 2.17 | 535,488 | 35.39 | 33.81–37.02 |
| 12 | Badakhshan | 3,109 | 3.56 | 1,130,535 | 27.50 | 26.54–28.48 |
| 13 | Parwan | 2,153 | 2.47 | 792,273 | 27.17 | 26.04–28.35 |
| 14 | Kabul | 14,695 | 16.84 | 5,966,395 | 24.63 | 24.23–25.03 |
| 15 | Sar-e-Pul | 1,148 | 1.32 | 666,737 | 17.22 | 16.24–18.24 |
| 16 | Faryab | 2,030 | 2.33 | 1,192,381 | 17.02 | 16.29–17.78 |
| 17 | Badghis | 681 | 0.78 | 575,212 | 11.84 | 10.97–12.76 |
| 18 | Wardak | 821 | 0.94 | 707,486 | 11.60 | 10.82–12.43 |
| 19 | Panjsher | 161 | 0.18 | 182,054 | 8.84 | 7.53–10.32 |
| 20 | Nuristan | 139 | 0.16 | 175,507 | 7.92 | 6.66–9.35 |
| 21 | Kunduz | 958 | 1.10 | 1,233,223 | 7.77 | 7.28–8.28 |
| 22 | Baghlan | 720 | 0.83 | 1,093,013 | 6.59 | 6.11–7.09 |
| 23 | Uruzgan | 234 | 0.27 | 467,659 | 5.00 | 4.38–5.69 |
| 24 | Takhar | 570 | 0.65 | 1,175,306 | 4.85 | 4.46–5.26 |
| 25 | Paktika | 211 | 0.24 | 830,994 | 2.54 | 2.21–2.91 |
| 26 | Zabul | 78 | 0.09 | 412,150 | 1.89 | 1.50–2.36 |
| 27 | Helmand | 281 | 0.32 | 1,552,838 | 1.81 | 1.60–2.03 |
| 28 | Paktia | 97 | 0.11 | 656,430 | 1.48 | 1.20–1.80 |
| 29 | Daykundi | 75 | 0.09 | 553,372 | 1.36 | 1.07–1.70 |
| 30 | Farah | 65 | 0.07 | 604,420 | 1.08 | 0.83–1.37 |
| 31 | Bamyan | 47 | 0.05 | 531,344 | 0.88 | 0.65–1.18 |
| 32 | Nimroz | 17 | 0.02 | 197,513 | 0.86 | 0.50–1.38 |
| 33 | Ghor | 51 | 0.06 | 833,304 | 0.61 | 0.46–0.80 |
| 34 | Ghazni | 1 | 0.00 | 1,461,703 | 0.01 | 0.00–0.04 |

*Note.* Provinces are ranked by reported incidence, not by absolute case count. Rates use the province population estimates
provided in the source workbook. Confidence intervals are exact Poisson intervals.

The geographical pattern differed when absolute case numbers were considered. Kabul reported the largest number of cases, with 14,695 cases, representing 16.84% of the national total. Herat followed closely with 14,598 cases (16.73%), while Nangarhar reported 8,408 cases (9.64%), Kandahar 7,579 (8.69%), and Balkh 7,043 (8.07%). Together, these five provinces accounted for approximately 60.0% of all CL cases reported nationally. Thus, although Jowzjan had the highest incidence rate, Kabul and Herat contributed the greatest absolute numbers of cases because of their larger populations.

At the lower end of the distribution, Ghazni reported only one case, corresponding to a rate of 0.01 per 10,000 population. Ghor, Nimroz, Bamyan, Farah and Daykundi also had rates below 1.5 per 10,000. These very low values should be interpreted as reported surveillance rates, as they may reflect differences in healthcare access, diagnostic capacity or reporting completeness in addition to variation in disease occurrence.

### Distribution in urban areas

Considerable variation was also observed among urban centers. Shiberghan, the capital of Jowzjan Province, had the highest urban reported incidence, with 3,366 cases among an estimated population of 209,121, equivalent to 160.96 cases per 10,000 population. Fayzabad had the second-highest urban rate, at 120.60 per 10,000, followed by Asadabad at 110.91 and Pul-e-Alam at 82.00 per 10,000.

Other urban centers with comparatively high rates included Sar-e-Pul, Aybak, Herat and Mehtarlam. In contrast, the country’s larger metropolitan centers had lower rates despite contributing substantial numbers of cases. Kabul city reported 12,332 cases, corresponding to 23.99 per 10,000 population, whereas Kandahar city reported 2,164 cases, equivalent to 30.42 per 10,000. Herat city recorded 4,515 cases and a higher urban rate of 69.22 per 10,000. The contrast between absolute numbers and population-based rates illustrates the strongly focal nature of reported urban transmission.

### Distribution in rural districts

Several rural districts recorded considerably higher incidence rates than either their respective provincial averages or the major urban centers. Panjwai District in Kandahar Province had the highest reported rural incidence, with 3,301 cases among 105,475 residents, corresponding to 312.96 cases per 10,000 population. This was followed by Ghoryan District in Herat Province, with 2,657 cases and a rate of 240.93 per 10,000, and Hazrat-e-Sultan District in Samangan Province, with 1,175 cases and a rate of 234.51 per 10,000.

Other prominent rural foci included Chaharasyab in Kabul Province (221.34 per 10,000), Aqcha in Jowzjan (199.19), Nadir Shah Kot in Khost (197.81), Arghandab in Kandahar (188.89), Dawlatabad in Balkh (168.85), Baharak in Badakhshan (167.46), Fayzabad District in Jowzjan (155.79) and Dawlatabad in Faryab (154.32). The high rates were therefore not confined to a single part of the country but were distributed across several western, northern, eastern and southern provinces.

The ranking of rural districts also differed depending on whether incidence rates or absolute case counts were considered. For example, Enjil and Guzara districts in Herat reported 2,423 and 2,059 cases, respectively, but their larger population denominators resulted in lower incidence rates than those observed in smaller districts such as Hazrat-e-Sultan and Chaharasyab.

### Monthly distribution of reported cases

The monthly distribution of reported CL cases was uneven throughout 2024 (Figure 2). May recorded the highest monthly number, with 10,351 cases, representing 11.86% of the annual total. November had the second-highest number, with 9,885 cases (11.33%), followed by December with 8,866 (10.16%) and October with 8,304 (9.52%).

**Figure 2.**
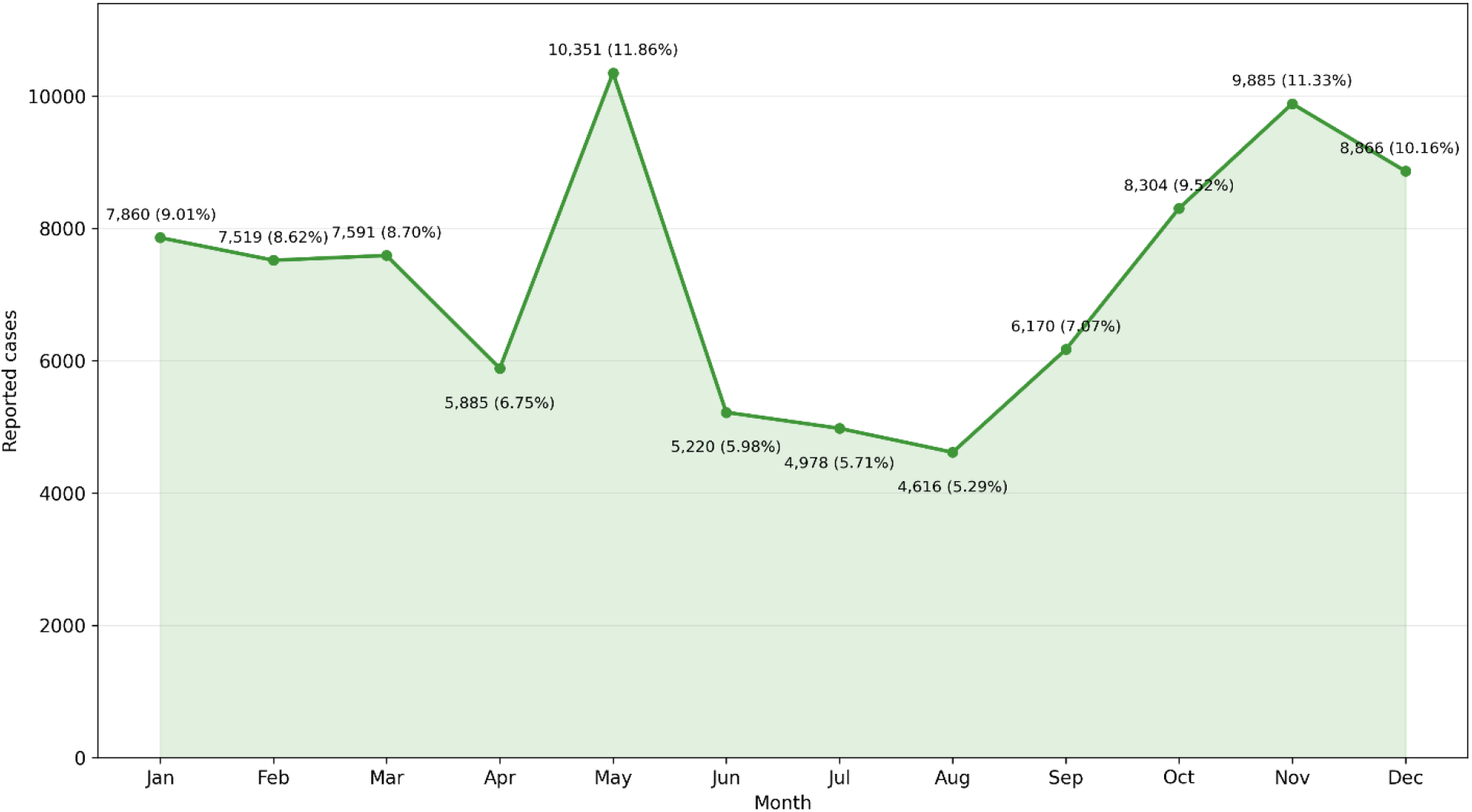
Monthly reported cutaneous leishmaniasis cases, Afghanistan, 2024.

The lowest monthly count occurred in August, when 4,616 cases were reported, accounting for 5.29% of all cases. July had the second-lowest count, with 4,978 cases (5.71%), followed by June with 5,220 (5.98%). After the peak in May, reported cases declined throughout the summer, reaching their lowest level in August, before increasing again from September and remaining relatively high through the end of the year. January, February, and March each accounted for between 8.6% and 9.0% of the annual total.

### Seasonal distribution

The corrected seasonal aggregation showed that autumn contributed the largest number of reported cases, with 24,359 cases, representing 27.92% of the annual total. Winter followed closely with 24,245 cases (27.79%), while spring accounted for 23,827 cases (27.31%). Summer had a substantially lower total of 14,814 cases, representing 16.98% of all reported cases (Table 3).

**Table 3.**
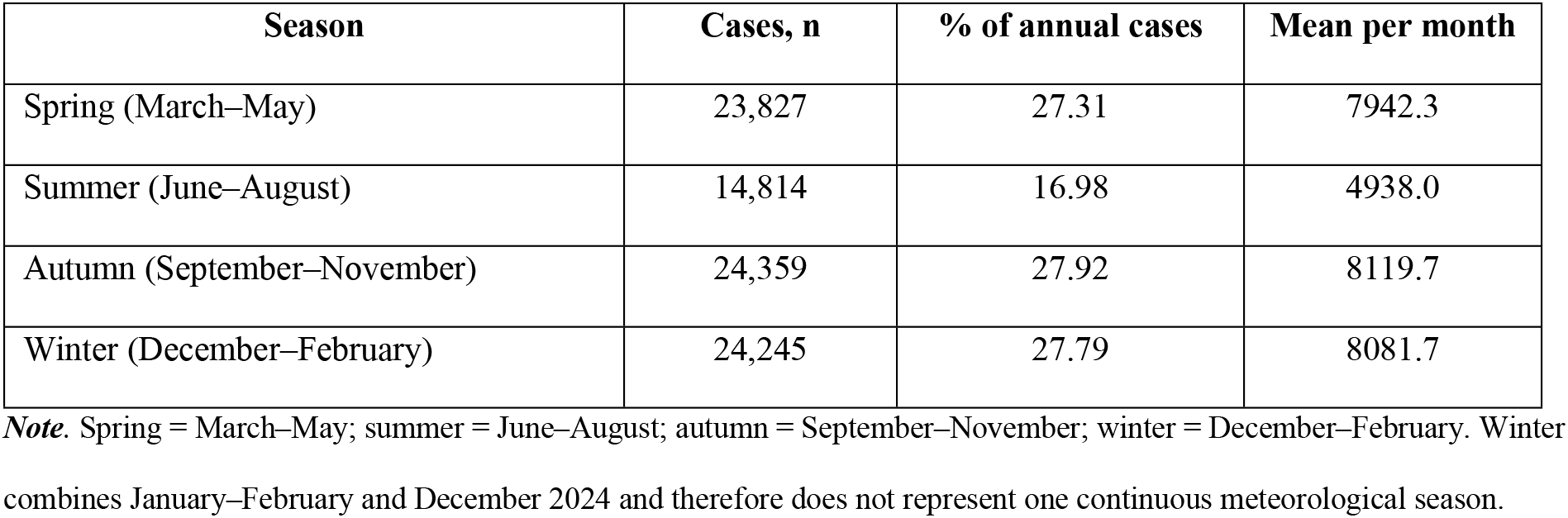
Seasonal distribution of reported cutaneous leishmaniasis cases.

The mean monthly case count was 8,119.7 during autumn, 8,081.7 during winter and 7,942.3 during spring, compared with 4,938.0 during summer. The temporal pattern therefore did not show a summer peak. Instead, reported cases were least frequent during June–August and increased during autumn, remaining high during the winter months. As the dataset covered only one calendar year, these findings describe the seasonal distribution observed in 2024 and should not be interpreted as evidence of a recurring long-term seasonal pattern.

### Distribution of zoonotic cutaneous leishmaniasis

All observations classified as zoonotic cutaneous leishmaniasis were recorded in Balkh Province. A total of 7,043 cases were reported across 14 districts and urban areas, corresponding to an overall reported incidence of 45.34 per 10,000 population. Dawlatabad recorded the highest district-specific incidence at 168.85 per 10,000 population, followed by Kaldar at 112.41 and Balkh District at 102.99 per 10,000. Mazar-e-Sharif contributed the largest absolute number of reported ZCL cases, with 1,538 cases, representing 21.84% of the provincial ZCL burden, followed by Balkh District with 1,507 cases (21.40%) and Dawlatabad with 1,390 cases (19.74%). No cases were reported from Charkent, Keshendeh or Zari. The district-level totals summed to 7,043 cases and 1,553,501 population, consistent with the programme-defined provincial denominator (Table 4).

**Table 4.** Corrected Distribution of Zoonotic Cutaneous Leishmaniasis in Balkh Districts.

| Rank | District | Cases, n | % of ZCL cases | Population | Rate per 10,000 | 95% CI |
| --- | --- | --- | --- | --- | --- | --- |
| 1 | Dawlatabad | 1,390 | 19.74 | 82,322 | 168.85 | 160.09–177.96 |
| 2 | Kaldar | 272 | 3.86 | 24,198 | 112.41 | 99.44–126.59 |
| 3 | Balkh | 1,507 | 21.40 | 146,318 | 102.99 | 97.86–108.33 |
| 4 | Charbulak | 790 | 11.22 | 98,074 | 80.55 | 75.03–86.37 |
| 5 | Khulm | 508 | 7.21 | 90,590 | 56.08 | 51.31–61.17 |
| 6 | Chemtal | 233 | 3.31 | 72,995 | 31.92 | 27.95–36.29 |
| 7 | Sholgareh | 427 | 6.06 | 138,837 | 30.76 | 27.91–33.82 |
| 8 | Nahr-e-Shahi | 154 | 2.19 | 54,375 | 28.32 | 24.03–33.16 |
| 9 | Mazar-e-Sharif | 1,538 | 21.84 | 550,291 | 27.95 | 26.57–29.38 |
| 10 | Shortepa | 115 | 1.63 | 47,969 | 23.97 | 19.79–28.78 |
| 11 | Dehdadi | 109 | 1.55 | 81,704 | 13.34 | 10.95–16.09 |
| 12 | Charkent | 0 | 0.00 | 53,805 | 0.00 | 0.00–0.69 |
| 13 | Keshendeh | 0 | 0.00 | 58,929 | 0.00 | 0.00–0.63 |
| 14 | Zari | 0 | 0.00 | 53,094 | 0.00 | 0.00–0.69 |
| **Total** |  | 7,043 | 100.00 | 1,553,501 | 45.34 | 44.28–46.41 |

*Note.* Districts are ranked by reported incidence per 10,000 population. The total row is not ranked. The erroneous nan row was removed, and Mazar-e-Sharif was restored. CI = confidence interval.

### Data completeness

The district-level dataset contained 363 records and 4,356 district–month cells. Of these, 1,763 cells (40.5%) were blank, and 70 district records contained no positive monthly count. The proportion of blank cells ranged from 33.06% in January to 46.28% in July. The source workbook did not allow blank cells to be reliably distinguished as true zero reports or missing and non-submitted reports. Consequently, the findings represent reported surveillance measures and may underestimate the underlying burden of CL, particularly in areas with incomplete reporting or limited access to diagnostic services.

The provincial subtotal rows were excluded from the district-level analysis to avoid double counting. Two duplicate province–district combinations were also identified; in both instances, the additional record contained no reported cases, and consolidation did not alter the national total. All recorded monthly case values were non-negative integers.

## Discussion

This nationwide ecological study demonstrated that cutaneous leishmaniasis remained a substantial and geographically heterogeneous public health problem in Afghanistan during 2024. A total of 87,245 cases were reported, corresponding to an overall reported incidence of 25.51 per 10,000 population. Anthroponotic cutaneous leishmaniasis accounted for 91.9% of the reported cases, whereas 8.1% were classified as zoonotic cutaneous leishmaniasis. Considerable variation was observed across provinces, urban centers and rural districts. Jowzjan had the highest provincial incidence, while Kabul and Herat contributed the largest absolute numbers of cases. Several localized urban and rural areas, particularly Shiberghan, Panjwai, Ghoryan and Hazrat-e-Sultan, recorded exceptionally high reported rates. Temporally, May had the highest monthly number of reported cases, while August had the lowest, and the seasonal burden was lowest during summer.

The substantial national burden observed in this study is consistent with Afghanistan’s historical recognition as one of the countries most severely affected by cutaneous leishmaniasis. An earlier investigation in Kabul estimated approximately 67,500 active cases during 2002–2003 and documented evidence of previous infection in a large proportion of residents in some districts [17]. Although direct comparison is difficult because the earlier study focused on Kabul and used different epidemiological methods, the present findings indicate that CL remains widely distributed beyond the historically recognized urban focus of the capital. Studies conducted in northern Afghanistan have also demonstrated the coexistence of anthroponotic and zoonotic transmission patterns and have emphasized differences in their transmission seasons and ecological characteristics [22,23]. The current nationwide findings extend this earlier evidence by showing that the reported burden is distributed across multiple northern, western, eastern, central and southern provinces.

The highly uneven provincial distribution reflects the focal epidemiology of CL. Jowzjan had the highest reported provincial incidence, at 104.99 cases per 10,000 population, and this elevated rate was strongly influenced by the burden recorded in Shiberghan. The concentration of cases in an urban center is epidemiologically compatible with anthroponotic transmission, which is commonly associated with densely populated communities in which infected humans may serve as the principal reservoir [2]. Housing conditions such as cracked mud walls, overcrowding, limited environmental sanitation and proximity between human dwellings and sandfly breeding or resting sites may increase human–vector contact. However, these factors were not measured in the present aggregated dataset. The high rate in Shiberghan should therefore be interpreted as an important epidemiological signal requiring household-level, environmental and entomological investigation rather than as confirmation of a specific transmission mechanism.

Kabul and Herat contributed the largest absolute numbers of cases, although their incidence rates were lower than that of Jowzjan. This difference illustrates the importance of distinguishing absolute burden from population-based incidence. Kabul’s large population means that even a moderate incidence rate can generate a substantial number of patients requiring diagnosis and treatment. Historical evidence has shown intense anthroponotic transmission in Kabul [17], and the continued high number of reported cases may reflect persistent transmission in densely populated urban communities. Population displacement, informal settlements, overcrowding and restricted access to timely treatment may also sustain transmission, particularly where untreated individuals remain infectious to sandfly vectors. Nevertheless, the present study did not contain information on housing, migration, treatment delays or individual exposure, and these explanations remain plausible rather than demonstrated.

Herat showed both a large absolute burden and a high provincial incidence. Several districts within the province, particularly Ghoryan, Enjil and Guzara, also contributed substantial case numbers or high population-based rates. Herat’s ecological diversity, movement between urban and rural areas, population mobility and proximity to endemic areas of neighboring Iran may be relevant to this pattern. Iran has reported marked geographical heterogeneity in CL, with differences according to parasite species, vector ecology, climate and settlement characteristics [10,11,13]. Cross-border movement could potentially contribute to the movement of infected individuals or exposure in endemic areas, but the surveillance data used in this study did not include travel histories, place of infection or parasite genotyping. The high burden in Herat should therefore support strengthened cross-border epidemiological awareness and targeted local investigation rather than assumptions regarding the origin of infection.

Kandahar also had a high provincial incidence, and Panjwai and Arghandab were among the most intensely affected rural districts. Panjwai recorded the highest district-level reported incidence in the country. Rural transmission may be influenced by housing construction, irrigation practices, agricultural activities, proximity to rodent habitats and greater exposure to sandflies during evening and night-time outdoor activities. Comparable associations with agricultural and outdoor exposure have been reported from other endemic settings, including Libya [9]. Nevertheless, high incidence in a rural district does not by itself establish zoonotic transmission. Confirmation would require parasite identification, vector studies and investigation of potential animal reservoirs. In the absence of these data, the high rates in Panjwai, Arghandab and other rural districts should be considered priorities for combined epidemiological, molecular and entomological studies.

The contrast between provincial averages and local district rates was particularly important. Ghoryan in Herat, Hazrat-e-Sultan in Samangan, Chaharasyab in Kabul, Aqcha in Jowzjan and Nadir Shah Kot in Khost recorded substantially higher rates than many provincial averages. This indicates that aggregation at the provincial level may conceal highly localized transmission foci. Similar spatial clustering has been described in Iran, where ecological and geographical conditions created distinct areas of elevated CL risk [10,11], and in Asir Province, Saudi Arabia, where cases were unevenly distributed across localities [16]. Targeting surveillance and control exclusively at the provincial level may therefore result in limited resources being distributed too broadly. District-level prioritization is likely to be more appropriate in locations with exceptionally high reported incidence.

The present study also showed that provinces with very low reported incidence, including Ghazni, Ghor, Nimroz, Bamyan and Farah, should not automatically be regarded as areas with little or no transmission. Routine passive surveillance is influenced by access to healthcare, clinical recognition, diagnostic capacity and reporting completeness. The finding that 40.5% of district–month cells were blank raises concern that some low rates may reflect incomplete or absent reporting rather than genuinely low disease occurrence. Conversely, areas with stronger diagnostic services, established treatment centers or referral facilities may report more cases because patients from neighboring locations seek care there. Differences in surveillance performance may therefore contribute to part of the observed geographical heterogeneity.

All cases classified as zoonotic cutaneous leishmaniasis in the source surveillance system were reported from Balkh Province. Within Balkh, Dawlatabad had the highest district-specific incidence, while Mazar-e-Sharif contributed the largest absolute number of ZCL cases. Earlier studies from northern Afghanistan demonstrated ecological and seasonal distinctions between anthroponotic and zoonotic CL and reported the effectiveness of integrated preventive interventions against zoonotic transmission [22,23]. The current concentration of program- classified ZCL in Balkh is therefore epidemiologically plausible. However, the surveillance classifications were not supported by patient-level molecular confirmation in the available dataset. The recorded categories should not be interpreted as definitive confirmation of *Leishmania major* or *Leishmania tropica* in every case. Molecular characterization of parasites, together with vector and reservoir studies, is needed to establish the transmission cycles operating in individual districts.

International comparisons also demonstrate that CL epidemiology varies considerably between settings. Studies from north-west Pakistan have reported substantial burdens among children and young people and seasonal increases during spring and summer [1]. Cases reported in Karachi have also been associated with travel from endemic areas, highlighting the potential influence of population movement on urban disease occurrence [15]. In contrast, studies from Iran have often documented increased clinical presentation during autumn, although patterns have varied across provinces and transmission settings [10,13]. Differences between the present findings and those from neighboring countries may be related to climatic conditions, sandfly species, parasite type, incubation period, healthcare-seeking behavior, diagnostic practices, study design, and the completeness of surveillance systems. These methodological and ecological differences limit direct comparison of reported incidence rates across countries.

The temporal distribution observed in Afghanistan did not show a summer peak. Reported cases declined after May, reached their lowest level in August, and increased again during autumn and winter. This pattern may partly reflect the interval between infection and clinical presentation.

CL lesions may become apparent weeks or months after an infective sandfly bite, and further delays may occur before patients seek care, receive a diagnosis and enter the surveillance system [2,5]. Consequently, the month of reporting may not correspond to the month of transmission.

Autumn increases have also been reported in parts of Iran and have been attributed to infections acquired during periods of greater sandfly activity becoming clinically apparent after the incubation period [10,13]. By contrast, the different seasonal pattern reported in north-west Pakistan illustrates that temporal distributions may vary according to local climate, vector ecology and surveillance practices [1].

Because the present analysis covered only one calendar year, the monthly and seasonal findings should be interpreted as descriptive temporal variation during 2024 rather than evidence of a stable recurring seasonal pattern. Reliable characterization of seasonality would require several consecutive years of surveillance data linked with dates of symptom onset, diagnostic confirmation, temperature, rainfall, humidity and local vector abundance. Nevertheless, the increase in reported cases during autumn and winter suggests that diagnostic and treatment services should remain sufficiently prepared during these periods. Vector-control activities should be timed according to local entomological evidence rather than solely according to the months in which clinical cases are reported.

The geographical concentration of the burden has important implications for public health planning. Kabul, Herat, Nangarhar, Kandahar and Balkh accounted for most reported cases, while Jowzjan and several individual districts had exceptionally high population-based rates. High-volume provinces require adequate diagnostic and treatment capacity because of their large caseloads, whereas smaller areas with very high incidence may benefit from intensive local surveillance, active case detection and operational investigation. Control strategies should also reflect the suspected transmission context. Prompt diagnosis and effective treatment are particularly important where anthroponotic transmission predominates, whereas confirmed zoonotic transmission may additionally require interventions directed at vectors, reservoirs and environmental conditions [2,22,23].

Overall, the study provides an updated national epidemiological baseline and demonstrates that the reported burden of CL in Afghanistan is both substantial and highly localized. The coexistence of high-burden urban centers and intensely affected rural districts indicates that a uniform national control strategy is unlikely to address the diversity of transmission settings.

Strengthening reporting completeness, standardizing diagnostic and surveillance procedures, documenting laboratory confirmation and conducting targeted molecular, entomological and environmental investigations in identified hotspots will be essential for designing locally appropriate and sustainable control interventions.

## Limitations

This study has several limitations. First, the analysis was based exclusively on cases reported through government health facilities and did not include patients diagnosed or treated in private clinics, non-governmental facilities or community-based services. The reported national burden is therefore likely to underestimate the true number of cutaneous leishmaniasis cases in Afghanistan. Second, the study relied on retrospectively collected routine surveillance data, which may have been affected by incomplete reporting, reporting delays and differences in diagnostic and surveillance capacity between geographical areas. Third, the aggregated nature of the dataset prevented assessment of individual-level risk factors, treatment outcomes and causal associations. Finally, because the analysis covered only one calendar year, the observed monthly and seasonal pattern should be interpreted as temporal variation during 2024 rather than as evidence of a stable long-term seasonal trend.

## Conclusion

Cutaneous leishmaniasis constituted a substantial and geographically uneven public health burden in Afghanistan during 2024, with 87,245 reported cases and an overall reported incidence of 25.51 per 10,000 population. The highest provincial incidence was observed in Jowzjan, followed by Herat and Kandahar, while several rural districts, particularly Panjwai, Ghoryan and Hazrat-e-Sultan, recorded exceptionally high rates. The temporal pattern did not show a summer peak; reported cases were lowest during summer and increased during autumn and winter, with May representing the highest individual monthly count. The coexistence of intense urban and rural foci points to a complex epidemiological pattern, although species-level confirmation is required before specific transmission cycles can be established. These findings support the prioritization of high-burden areas for strengthened surveillance, active case detection, timely diagnosis and treatment, and locally appropriate vector-control measures. Improvements in reporting completeness and diagnostic capacity are also essential to obtain a more accurate estimate of the national burden and to guide sustainable control strategies.

## Data Availability

The authors confirm that all data generated or analyzed during this study are included in this article. If any further information about the data is needed, we can provide it.

## Acknowledgements

We are heartily grateful to all the staff members of the Afghanistan National Malaria and Other Vector-Borne Diseases Program and Kandahar University for their cooperation and sincere assistance. We are thankful to Dr Bismillah Jamal, Dr Rohullah Jamal, Dr Hizbullah Fitrat, and Dr Abdul Basit Noorzai from the Ministry of Public Health for their kind support in providing the raw data. We are grateful to the officials of Kandahar University Faculty of Medicine for providing full support to conduct this research.

## Authors’ contributions

Conceptualization: BAR, WRT

Data curation: BAR, WML, AJS

Formal analysis: BAR, WML

Funding acquisition: No funding

Investigation: BAR

Methodology: BAR, WML, AJS, WRT

Project administration: BAR

Resources: BAR, WML, AJS, WRT

Software: BAR, WML

Supervision: BAR

Validation: BAR, WML, AJS, WRT

Visualization: BAR, WML, AJS, WRT

Writing – original draft: BAR

Writing – review & editing: BAR, WML, AJS, WRT

All authors have read and approved the manuscript.

